# A Mixed Methods System for the Assessment of Post Exertional Malaise in Encephalomyelitis/Chronic Fatigue Syndrome

**DOI:** 10.1101/2023.04.24.23288821

**Authors:** Barbara Stussman, Brice Calco, Gina Norato, Angelique Gavin, Snigdha Chigurupati, Avindra Nath, Brian Walitt

## Abstract

**Background:** A central feature of Myalgic Encephalomyelitis/Chronic Fatigue Syndrome (ME/CFS) is post exertional malaise (PEM), which is an acute worsening of symptoms after a physical, emotional and/or mental exertion. PEM is also a feature of Long COVID. Dynamic measures of PEM have historically included scaled questionnaires which have not been validated in ME/CFS. To enhance our understanding of PEM and how best to measure it, we conducted semi-structured qualitative interviews (QIs) at the same intervals as Visual Analog Scale (VAS) measures after a Cardiopulmonary Exercise Test (CPET).

**Methods:** Ten ME/CFS and nine healthy volunteers participated in a CPET. For each participant, PEM symptom VAS (7 symptoms) and semi-structured QIs were administered at six timepoints over 72 hours before and after a single CPET. QI data were used to plot the severity of PEM at each time point and identify the self-described most bothersome symptom for each patient. QI data were used to determine the symptom trajectory and peak of PEM. Performance of QI and VAS data were compared to each other using Spearman correlations.

**Results:** QIs documented that each ME/CFS volunteer had a unique PEM experience, with differences noted in the onset, severity, trajectory over time, and most bothersome symptom. No healthy volunteers experienced PEM. Scaled QI data were able to identify PEM peaks and trajectories, even when VAS scales were unable to do so due to known ceiling and floor effects. QI and VAS fatigue data corresponded well prior to exercise (baseline, r=0.7) but poorly at peak PEM (r=0.28) and with the change from baseline to peak (r=0.20). When the most bothersome symptom identified from QIs was used, these correlations improved (r=.0.77, 0.42. and 0.54 respectively) and reduced the observed VAS scale ceiling and floor effects.

**Conclusion:** QIs were able to capture changes in PEM severity and symptom quality over time in all the ME/CFS volunteers, even when VAS scales failed to do so. Information collected from QIs also improved the performance of VAS. Measurement of PEM can be improved by using a quantitative-qualitative mixed model approach.

**Disclaimer:** This research/work/investigator was supported (in part) by the Division of Intramural Research of the National Institutes of Health, NINDS. The content is solely the responsibility of the author(s) and does not necessarily represent the official views of the National Institutes of Health.

## INTRODUCTION

Persistent and disabling fatigue, exercise intolerance, cognitive difficulties, and myalgias/arthralgias are characteristic of a medical disorder referred to as Myalgic Encephalomyelitis/Chronic Fatigue Syndrome (ME/CFS). Post exertional malaise (PEM) is an acute worsening of these symptoms after minimal physical or mental exertion (1). PEM patients describe the experience as all-encompassing with symptoms affecting every part of the body, difficult to predict or manage, and requiring complete bedrest to recover. More recently, evidence has emerged of PEM in persons with long COVID (2,3). While PEM has a wide range of symptoms, three core symptoms have been identified: exhaustion, cognitive difficulties, and neuromuscular complaints (4). Although PEM is considered a central feature of ME/CFS (5) its assessment is challenging due to its subjective nature and a lack of validated tools for reliable quantification. Cardiopulmonary exercise testing (CPET) is an important tool for the evaluation of patients with ME/CFS (5,6) and is used to induce PEM in research settings. The quality and depth of severity of PEM experienced by patients after undergoing CPET can be gleaned from the words of one of our ME/CFS patients: “[After CPET] I go into this shutdown mode. I am aware of what’s going on around me…but I need to save energy. So I’ll point to things and if I need to say some words, I’ll say them. I laid down right away… Everybody was doing all sorts of things around me…and I just didn’t talk.” The development of a scale that can accurately and reliably capture how this severe symptomatic experience evolves over time is an essential first step to understanding the biology of PEM.

Historically, Visual Analogue Scales (VAS) have been widely used for monitoring the course of chronic diseases and capturing changes due to medical interventions and have been validated for detecting pain in several patient populations (7-9). In the ME/CFS patient population, VAS and Numeric Rating Scales (NRS) have been employed to detect differences in physical fatigue, mental fatigue, and painful symptoms between patients and healthy controls following stress imposed by exercise or orthostatic testing (10-16). Current scales are known to produce ceiling effects and are sensitive to minor variability in wording when measuring PEM (17-19). Further complicating measurement is the characteristically delayed onset of PEM. PEM in ME/CFS patients following CPET peaks within hours to days afterward with a duration of several days or longer (4, 20, 21). Measuring the evolution of PEM requires point-in-time measurements, which have been performed to date with VAS and NRS (6, 22, 23). Both of these types of measurement tools are psychometric response scales used to measure symptom severity at a point-in-time in individual patients and have been shown to be highly correlated for such measurement (24-26). Neither type has been validated for measurement of PEM.

While the recently developed DePaul Symptom Questionnaire (DSQ) has provided the field a validated tool for assessing PEM in ME/CFS patients (17, 19, 27), the instrument uses lengthy recall periods and was not designed to capture PEM in real time. A variety of other retrospective questionnaires have been used to capture PEM symptom breadth, severity, and duration, such as the Patient-Reported Outcomes Measurement Information System (PROMIS), Short Form 36 Health Survey Questionnaire (SF-36), Chronic Fatigue Questionnaire (CFQ), and Fatigue Impact Scale (FIS) (19, 28, 29). Many of these instruments use lengthy temporal intervals in symptom assessment, such as seven days, thirty days, or six months, and thus are unsuitable for measuring moment-to-moment changes in PEM.

Open-ended questionnaires have been used to capture PEM following CPET. For instance, Twomey et al. (30) used a shorter recall period by providing participants an open-ended questionnaire 96 hours after exercise testing with instructions to recall the previous four days. Other studies have used open-ended questionnaires to collect point-in-time measurement of PEM following exercise testing by providing the questionnaires ahead of time and instructing patients to answer questions at several timepoints following the exercise test (11, 31). These types of retrospective methodologies and the use of predetermined questions are unavoidably subject to recall bias and cannot fully capture the individualistic nature and complexity of PEM (4, 17, 18, 20).

An interactive assessment allowing for probing and clarifying the breadth and severity of symptoms at a point-in-time during the experimental initiation of PEM is an important step toward the discovery of biological correlates. Qualitative interviews (QIs) afford patients the opportunity to fully delineate the breadth and complexity of the experience of PEM without being confined to singular questions as they are experiencing it. The measurement of fatigue is often used as a surrogate measure of PEM because fatigue is the most frequently reported PEM symptom (4, 20, 31). However, the experience of PEM is uniquely personal and is not uniformly defined by fatigue. PEM may be better measured if the most bothersome symptom for each person is considered when determining its severity. The current study is the first to use QIs to capture PEM symptoms at structured intervals following CPET testing. The current study aims to improve current measurement of PEM by employing a mixed-methods approach (collection and analysis of both quantitative VAS and qualitative QI data) to concurrently measure PEM in real-time. In this study, we evaluate the benefits and drawbacks of this method, determine if the method improves measurement performance over the standard use of PEM VAS scales, and make this approach available to other interested clinicians and scientists in the field.

## MATERIALS AND METHODS

### Study design

Data were collected as part of the Myalgic Encephalomyelitis/Chronic Fatigue Syndrome Protocol at the National Institutes of Health (NCT02669212), which was approved by the NIH Institutional Review Board (IRB). This was a deep phenotyping study of ME/CFS and healthy volunteers (HVs) that included a CPET intervention designed to induce PEM with serial follow-up performed over 72 hours. The current study used a convergent mixed methods study design in which quantitative and qualitative data were collected simultaneously; the data were analyzed independently and then merged and interpreted together (32). We chose this method to allow for a nuanced understanding of PEM, deemed imperative due to the wide variation in PEM between individuals found in our previous focus group study (4). The objective of the current study was to use qualitative data to improve upon the performance of standard VAS measurements in determining the peak time that PEM occurs and in determining relative severity over time within an individual experience of PEM.

### Participants

Study recruitment occurred between December 2016 and February 2020. Of 484 ME/CFS inquiries for NCT02669212, 217 individuals underwent detailed case reviews, 27 ME/CFS and 25 HVs underwent in-person research evaluation. Of these, a subgroup of 10 participants with ME/CFS and 9 HVs completed the CPET experiment. All ME/CFS participants met 2015 IOM ME/CFS criteria and were determined to have ME/CFS by a panel of clinical experts by unanimous consensus. Additional recruitment was terminated due to the COVID-19 pandemic. The study was approved by the NIH IRB. Informed consent was obtained from all participants.

### Cardiopulmonary Exercise Testing (CPET)

CPET is an exercise physiology protocol that is typically used to measure exercise performance and tolerance. It typically involves performing exercise on a cycle ergometer that starts at a level considered easy and steadily becomes more challenging over time during the same session.

Participants are instructed to exercise until they reach subjective exhaustion and cannot continue to exercise further (33). Small clinical studies report that a single CPET session (1-day CPET) is a reliable way to induce PEM in participants with ME/CFS (11, 34). The NIH protocol used single session CPET as a method to induce PEM for scientific inquiry. All volunteers rested for at least two days prior to undergoing CPET and met the criteria for a successful aerobic effort on CPET, with respiratory exchange ratio of 1.1 or greater.

### Data collection

Based on our previous research that confirmed the value of capturing textual data and multiple timepoints post CPET in assessing PEM (4), semi-structured QIs were conducted at several timepoints before and after volunteers underwent CPET testing (one hour pre-CPET; one, four, 24, 48 and 72 hours post-CPET). By performing an assessment before undergoing CPET, we established each volunteer’s pre-CPET baseline. We sought to fully understand physical, cognitive, and emotional symptoms experienced by both ME/CFS and HVs following CPET and to ascertain perceptions of the changes in symptom severity between timepoints. ME/CFS volunteers were asked to report their most bothersome symptom at each timepoint and to use examples from their daily lives as a benchmark to clarify the magnitude of PEM they were experiencing. Interviews were conducted in-person in the volunteer’s hospital room by an experienced qualitative researcher, with the exception of the four hour post CPET interview during which volunteers were enclosed in a metabolic measurement room; these interviews were conducted over the telephone. All interviews were audio recorded and transcribed by a professional service. Interview questions are shown in Table 1.

**Table 1.**
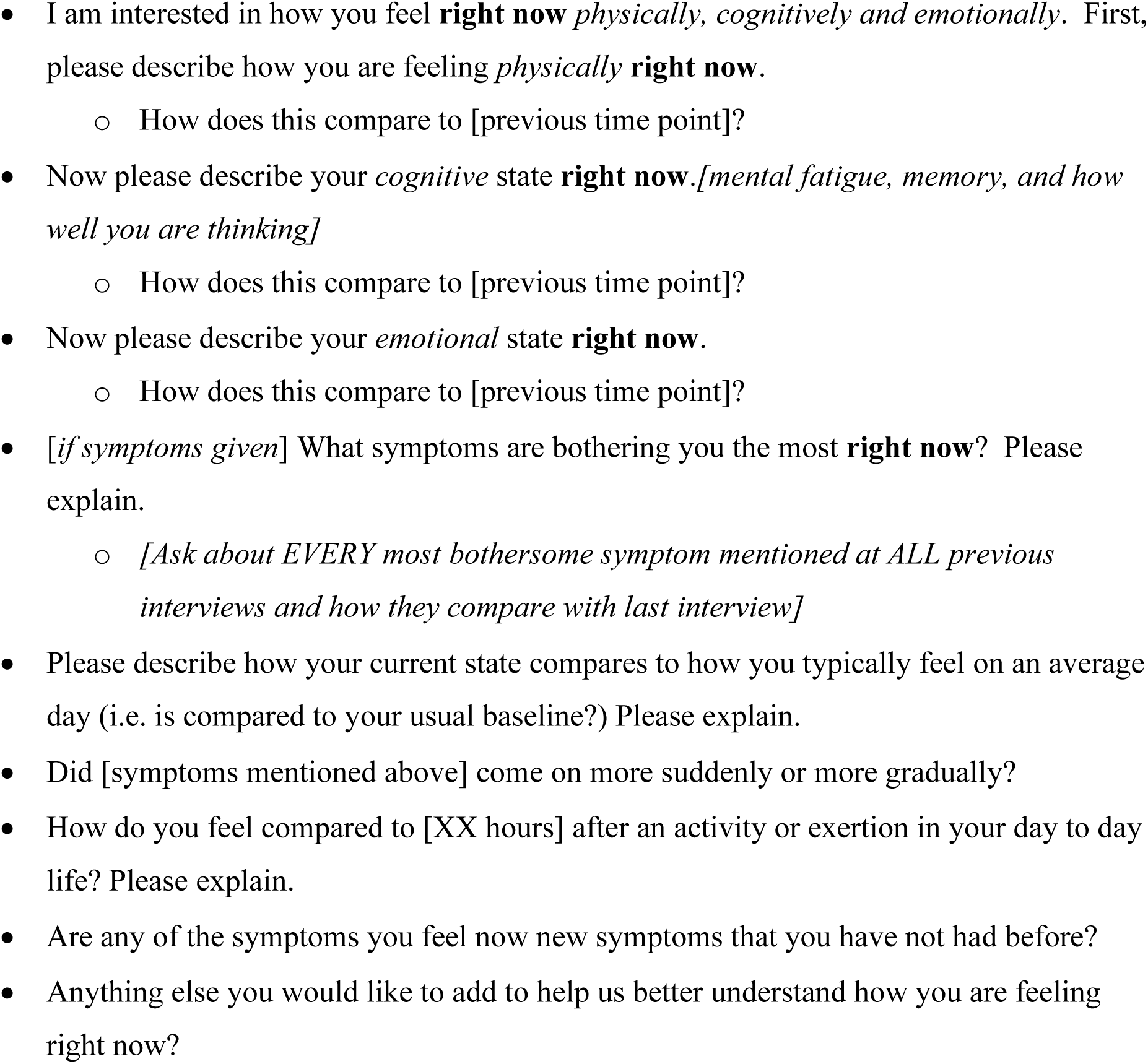
Qualitative Interview questions asked at 6 time points before and following CPET.

VAS data were collected for seven symptoms immediately following the QI and at the same six timepoints. These symptoms were: physical fatigue, mental fatigue, muscle aches, joint aches, headache, muscle weakness, and lightheadedness. Volunteers were provided a tablet and instructed “to place an “X” on a line for each symptom to indicate how they felt RIGHT NOW.” (Figure 1). Lines were anchored on the left and right side indicating “NOT AT ALL” and “MOST EXTREME.”

**Figure 1.**
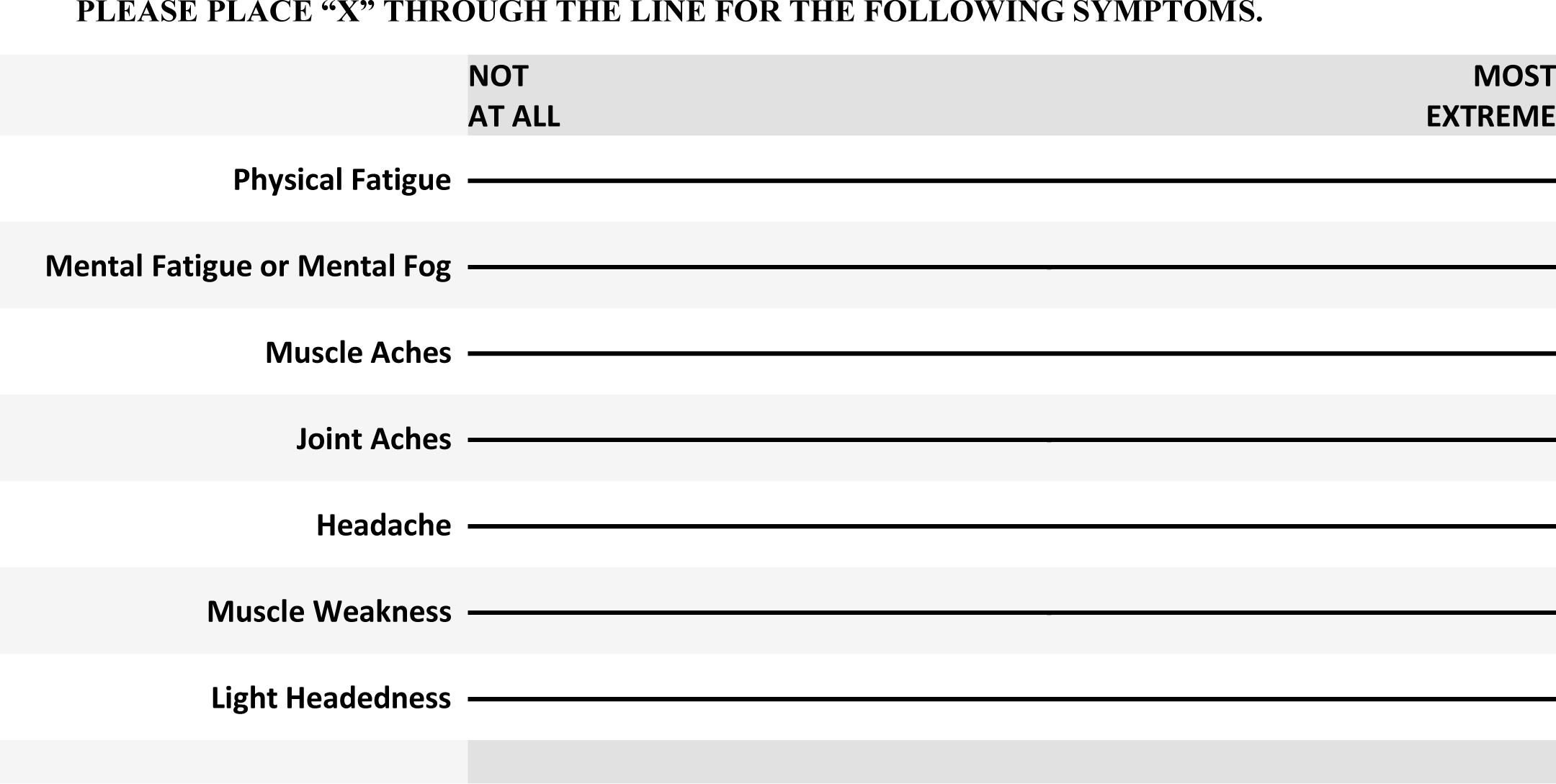
Visual analogue Scale (VAS) for PEM Symptoms. DIRECTIONS: You are asked to place an “X” through these lines to indicate how you are feeling RIGHT NOW. The left end of the line represents feeling good (experiencing no symptoms), while the right end of the line represents feeling your worst (experiencing your most severe symptoms).

Below is an excerpt from an interview at four hours post CPET that illustrates how the QI can provide contextual information to aid in the interpretation of VAS scores. In the example, the ME/CFS volunteer described the change in their headache from one hour post-CPET to four hours post-CPET and the patient’s VAS score for headache was 7.8 at one hour post-CPET and 9.6 at four hours post-CPET.

***Q: So can you tell me how you’re feeling right now physically?***

***A: Still fairly exhausted. And I have a headache not much different than what I felt like when I first came out [of CPET]. A little bit better, but not much***.

***Q: What symptom is bothering you the most right now?***

***A: Probably my headache is the most stressing part right now. Q: Is it as bad as when we spoke 3 hours ago?***

***A: It’s probably one degree less. It’s probably at a 7 instead of an 8/9 or 9/10. It’s a 7/8***.

This example highlights how the QI and VAS can describe different stories. Since the VAS is captured in isolation at each timepoint, the detection of change between timepoints is vulnerable to subjective interpretation of the scale in the moment with no ability to directly ask about differences between points. Furthermore, headache, not fatigue, was the most bothersome symptom. Asking volunteers to compare to the previous timepoint enabled researchers to graphically plot the course of PEM throughout the six timepoints.

## Data analysis

### Qualitative Interviews

Qualitative data analysis was performed on QI transcripts from all timepoints for each volunteer. Four researchers individually read and analyzed hundreds of pages of transcripts, evaluated, and synthesized the entirety of the PEM experience as described by patients. Each researcher separately plotted the trajectory of PEM across the six timepoints and individually determined the time of peak PEM and the most bothersome symptom. Examples of triggering events in their daily lives provided by ME/CFS volunteers served as benchmarks when rating the severity of PEM. Based on these benchmarks, six categories of severity were created: usual baseline (how participant usually feels on a typical day), slightly worse than usual baseline, somewhat worse than usual baseline, much worse than usual baseline, equivalent to severe triggering event, worse than severe triggering event. To meet the threshold on QI for having PEM, both an increase in PEM from the baseline value and a peak rating of three (much worse than baseline) or greater was required. Figure 2 provides an example of PEM following CPET using the QI scale. The most bothersome symptom was determined by analyzing each ME/CFS volunteer’s self-described experience of symptoms at each timepoint. The research team held in-depth consensus meetings to compare results and adjudicate any disagreement in the findings from the independent analyses. Before adjudication, the team reached 75% agreement based on their independent analyses. Complete consensus was achieved by the study team reviewing transcripts line-by-line together with in-depth discussions.

**Figure 2.**
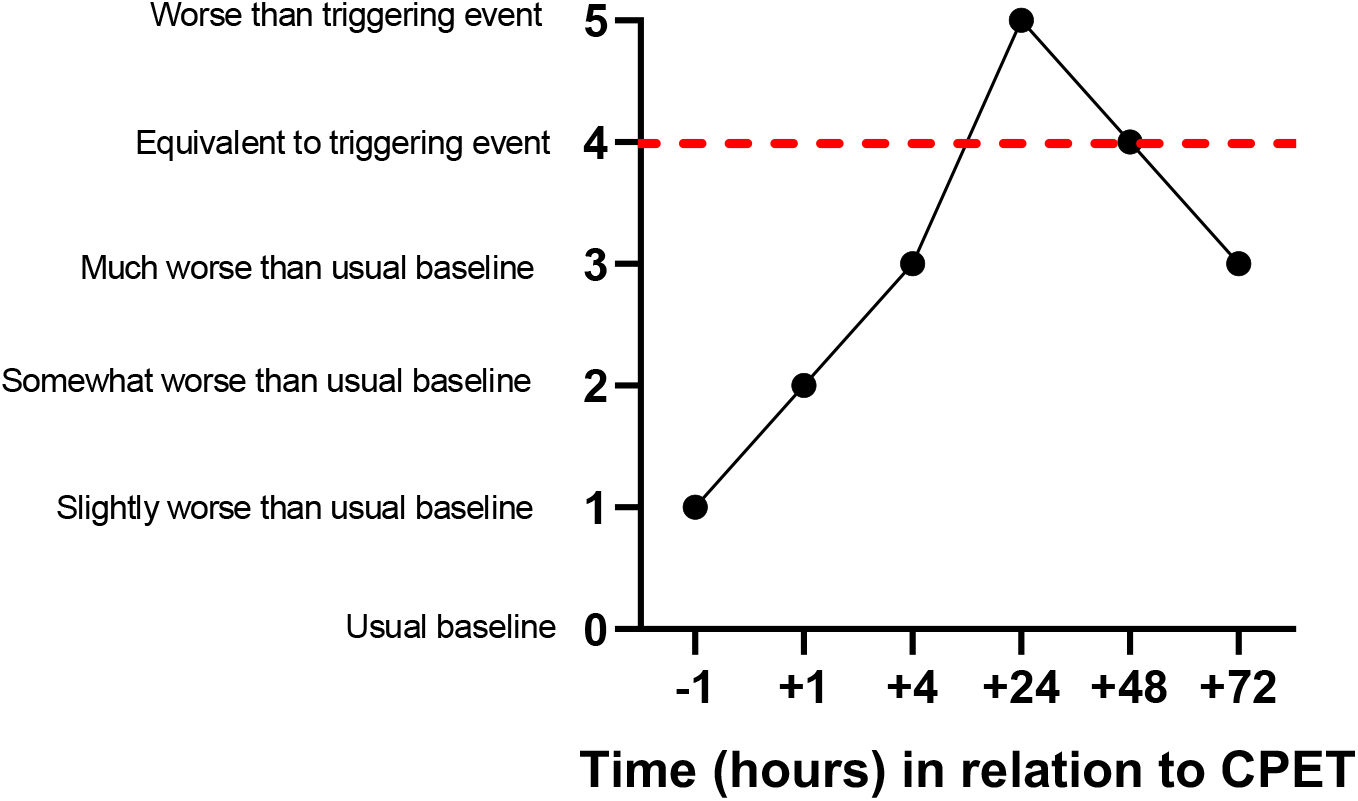
PEM Scaling Example Based on a Qualitative Interview. HVs experienced none or minor symptoms following CPET and the minor variance in symptoms aligned closely with the VAS rating for physical fatigue (Figure 3), suggesting HVs equated their post-CPET symptoms with physical fatigue. Because HVs did not experience PEM, the remainder of this paper focuses on ME/CFS volunteers.

### Quantitative data

PEM data, both derived from QIs and VAS, was investigated graphically within each volunteer to understand the time courses from pre-CPET to 72 hours post-CPET. Since all the ME/CFS and none of the HVs developed PEM, no comparisons were performed between the groups.

Correlations between QI and VAS data were performed to inform the utility of using QIs in conjunction with VAS in capturing PEM in ME/CFS volunteers. Correlations between PEM rating and VAS scores were visualized and tested at both baseline and at time of peak PEM. Longitudinal time courses were also investigated for the severity of physical fatigue, mental fatigue, headache, and muscle ache symptoms collected with VAS scales. These four symptoms encompassed the VAS categories that patients described as their most bothersome symptom. For each participant, correlations were also conducted to describe the relationship between QI PEM and physical fatigue VAS severity ratings. These correlations were repeated using each patient’s most bothersome symptom VAS severity rating. Change in symptoms from baseline to peak PEM was determined by subtracting the baseline from the maximum PEM score. Spearman correlations were used throughout. R version 4.0.0 was used for data analysis and visualization (35). Correlation coefficients whose magnitude was >0.68 are strong/highly correlated, 0.36-0.67 considered modest/moderately correlated, and <0.36 as low/weakly correlated (36).

## RESULTS

Demographic characteristics for study volunteers are shown in Table 2. Figure 4 shows the course of PEM symptom severity for ME/CFS volunteers across the six timepoints overlayed with Physical Fatigue VAS. QI data revealed that every ME/CFS volunteer experienced PEM within the 72-hour study period with only one returning to their pre-CPET level by the final 72-hour timepoint. A wide variation was seen in timing of peak PEM with occurrences at every timepoint measured after CPET. When comparing QI severity to VAS severity data, physical fatigue VAS failed to capture PEM for 30% of ME/CFS volunteers (7, 11, and 12) with the VAS plot line flat throughout the time course. Volunteer 5 shows a potential confounding issue with QIs, as the baseline value was rated a 4 (equivalent to severe triggering event). However, this volunteer rated their symptoms a 5 (worse than severe triggering event) at 24 hours suggestive of post-CPET PEM.

**Table 2.** Demographic characteristics for ME/CFS patients and Healthy Volunteers.

|  | ME/CFS<br>Patients (n=10) | Healthy<br>volunteers (n=9) |
| --- | --- | --- |
| **Sex** |  |  |
| Male | 5 | 6 |
| Female | 5 | 3 |
| **Age** |  |  |
| 18-29 | 3 | 2 |
| 30-39 | 1 | 2 |
| 40-49 | 4 | 2 |
| 50-59 | 1 | 3 |
| 60+ | 1 | -- |
| **Race** |  |  |
| White | 8 | 8 |
| Non-White | 2 | 1 |
| **Ethnicity** |  |  |
| Hispanic | 1 | -- |
| Non-Hispanic | 9 | 9 |
| **Geographic Region\*** |  |  |
| Northeast U.S. | 2 | -- |
| Southern U.S. | 2 | 9 |
| Midwestern U.S. | 1 | -- |
| Western U.S. | 4 | -- |
| Canada | 1 | -- |
| **Employment Status** |  |  |
| Full-time | -- | 5 |
| Part-time | 4 | 3 |
| Student | 1 | 1 |
| Not working due to disability | 4 | -- |
| Not working for other reasons | 1 | -- |
| **Marital Status** |  |  |
| Married | 7 | 1 |
| Divorced | -- | 2 |
| Never Married | 3 | 6 |
| **Education** |  |  |
| Less than college degree | 3 | 3 |
| College degree | 3 | 2 |
| Graduate level or above | 4 | 4 |
| **Years since symptom onset** |  |  |
| 1-2 | 4 | N/A |
| 3-4 | 1 | N/A |
| 5-6 | 5 | N/A |

\*Based on U.S. Census Bureau groupings

**Figure 3:**
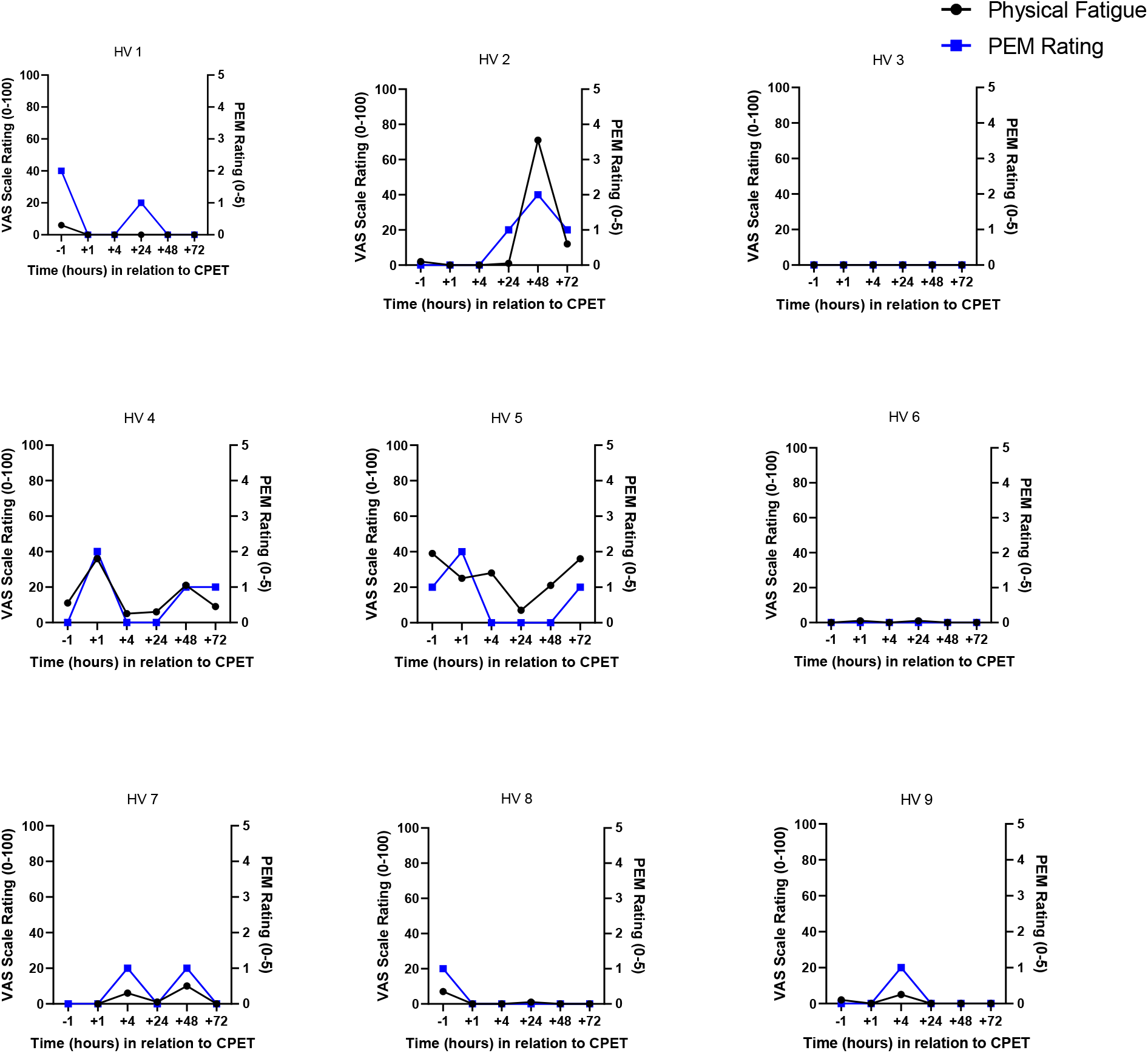
Overlay of PEM and Physical Fatigue VAS for Healthy Volunteers.

**Figure 4.**
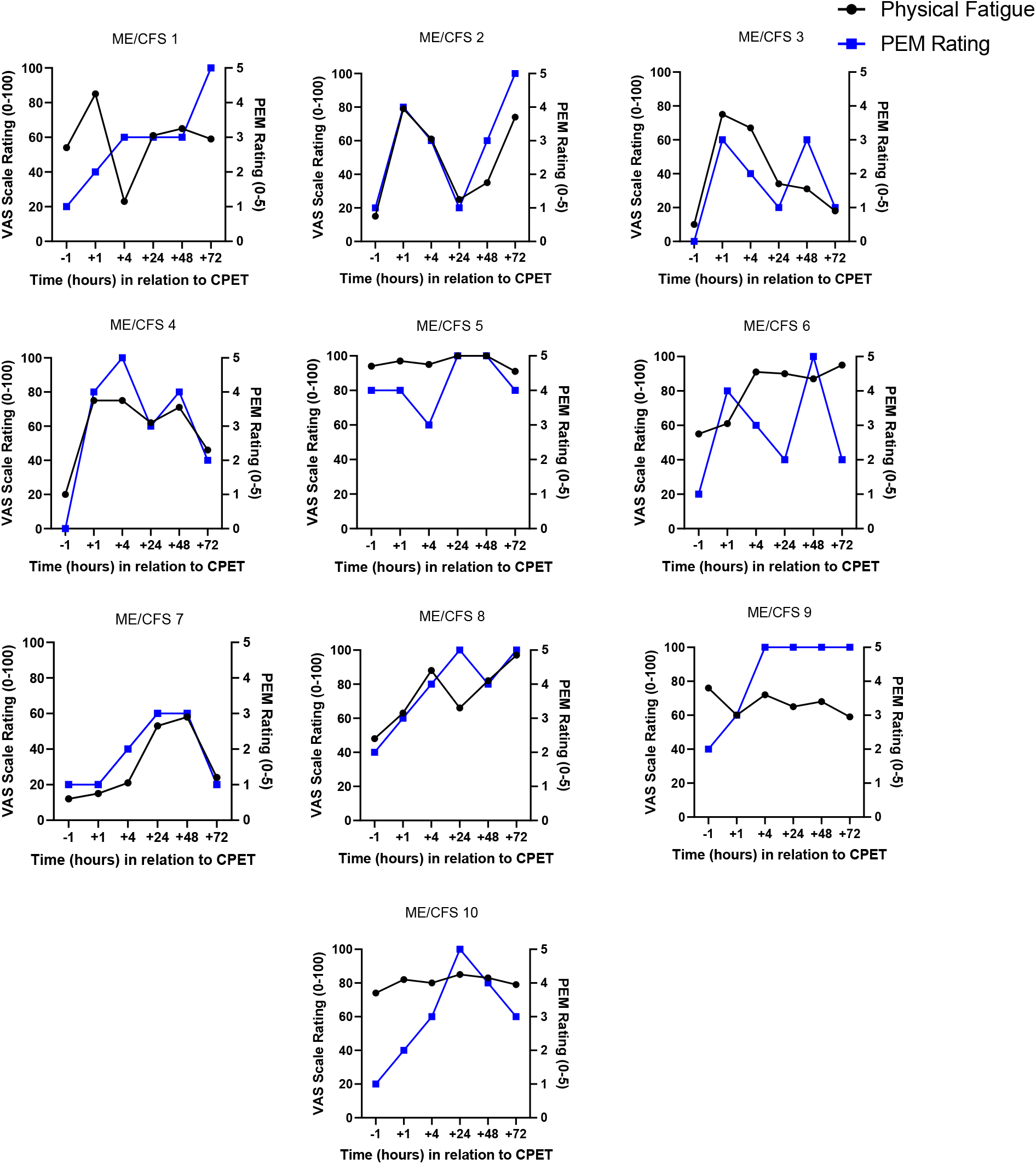
Overlay of PEM and Fatigue VAS for ME/CFS Patients.

For half of the ME/CFS volunteers, PEM severity and VAS physical fatigue severity aligned closely indicating that in these cases, the physical fatigue VAS captured PEM. In addition, several volunteers reported having more than one PEM peak during the time course on QIs, which was not captured by the physical fatigue VAS. Taken as a whole, these data show that interviews and VAS provided different information about the peak and course of PEM, with QIs having more measurement granularity and face validity.

QI data analysis revealed four symptoms as most bothersome for ME/CFS volunteers: physical fatigue (40%), mental fatigue (20%), headache (30%), and muscle ache (10%). Table 3 presents r values for the correlations of PEM between the QI and VAS severity scores when measured at baseline, at the time of peak PEM, as well as the change between baseline and peak PEM for the four most bothersome symptoms. When looking at the data combined across volunteers, singling out the most bothersome symptom outperforms the fixed VAS scales for all four symptoms at all three timepoints. When focusing on individual timepoints, strong correlations were seen for physical fatigue (r = 0.70) and mental fatigue (r = 0.66) at baseline, whereas muscle ache and headache had weak correlations (r = 0.39 and 0.34 respectively). However, at the time of peak PEM, the same correlations were uniformly weak. When change over time from baseline to peak PEM was considered, correlations between QIs and VAS were also weak (r = 0.03 to 0.37).

**Table 3.** R Values for Correlations Between PEM and VAS Among ME/CFS Volunteer’s Report of Most Bothersome Symptom (n=10)

|  | **ME/CFS Volunteers' Most Bothersome Symptom** |  |  |  |  |
| --- | --- | --- | --- | --- | --- |
|  | **Combined Most Bothersome Symptom for All Volunteers (n=10)**<br>**r (p value)** | **Physical Fatigue (n=4)**<br>**r (p value)** | **Mental Fatigue (n=2)**<br>**r (p value)** | **Muscle Ache (n=1)**<br>**r (p value)** | **Headache (n=3)**<br>**r (p value)** |
|  | **Correlations for Selected Timepoints for All ME/CFS Volunteers Combined** |  |  |  |  |
| Baseline | 0.77 (0.01) | 0.70 (0.02) | 0.66 (0.04) | 0.39 (0.27) | 0.34 (0.34) |
| Peak | 0.42 (0.23) | 0.28 (0.43) | 0.29 (0.42) | 0.28 (0.43) | 0.05 (0.89) |
| Change from Baseline to Peak | 0.54 (0.11) | 0.20 (0.57) | 0.03 (0.94) | 0.23 (0.53) | 0.37 (0.3) |
|  | **Correlations Across All Timepoints for Individual ME/CFS Volunteers** |  |  |  |  |
| ME/CFS 1 |  |  |  |  | 0.09 (0.86) |
| ME/CFS 2 |  | 0.35 (0.49) |  |  |  |
| ME/CFS 3 |  |  | 0.71 (0.12) |  |  |
| ME/CFS 4 |  |  | 0.96 (0.003) |  |  |
| ME/CFS 5 |  |  |  |  | 0.94 (0.005) |
| ME/CFS 6 |  |  |  |  | 0.70 (0.12) |
| ME/CFS 7 |  | 0.75 (0.08) |  |  |  |
| ME/CFS 8 |  | 0.74 (0.1) |  |  |  |
| ME/CFS 9 |  |  |  | 0.34 (0.51) |  |
| ME/CFS 10 |  | 0.90 (0.01) |  |  |  |

Table 3 also presents correlations between PEM and VAS severity at the individual level across all timepoints. For seven of the ten ME/CFS volunteers, the correlation between VAS and QI severity data was at or above .70 (r = 0.7 to 0.96). For the other three, the correlations were weak (r = 0.09 to 0.35). Due to these strong correlations and that these four symptoms are the most common PEM symptoms reported by ME/CFS patients (37, 20, 4), the remaining analyses describe correlations for only these four symptoms.

Figure 5 shows VAS severity across the six timepoints for the four most bothersome symptoms among ME/CFS volunteers. A wide variation was seen, with several scores sustaining high or low levels throughout the six timepoints, indicative of VAS ceiling (red) and floor (blue) effects. Additionally, of all the floor or ceiling effects, only one represented a ME/CFS volunteer’s most bothersome symptom (headache, red dashed line). By collecting information about the most bothersome symptom, the QIs complement the VAS scales, enabling selection of the most meaningful VAS measure for measuring PEM. However, QIs on their own performed better than the most bothersome symptom VAS in determining the occurrence of PEM.

**Figure 5.**
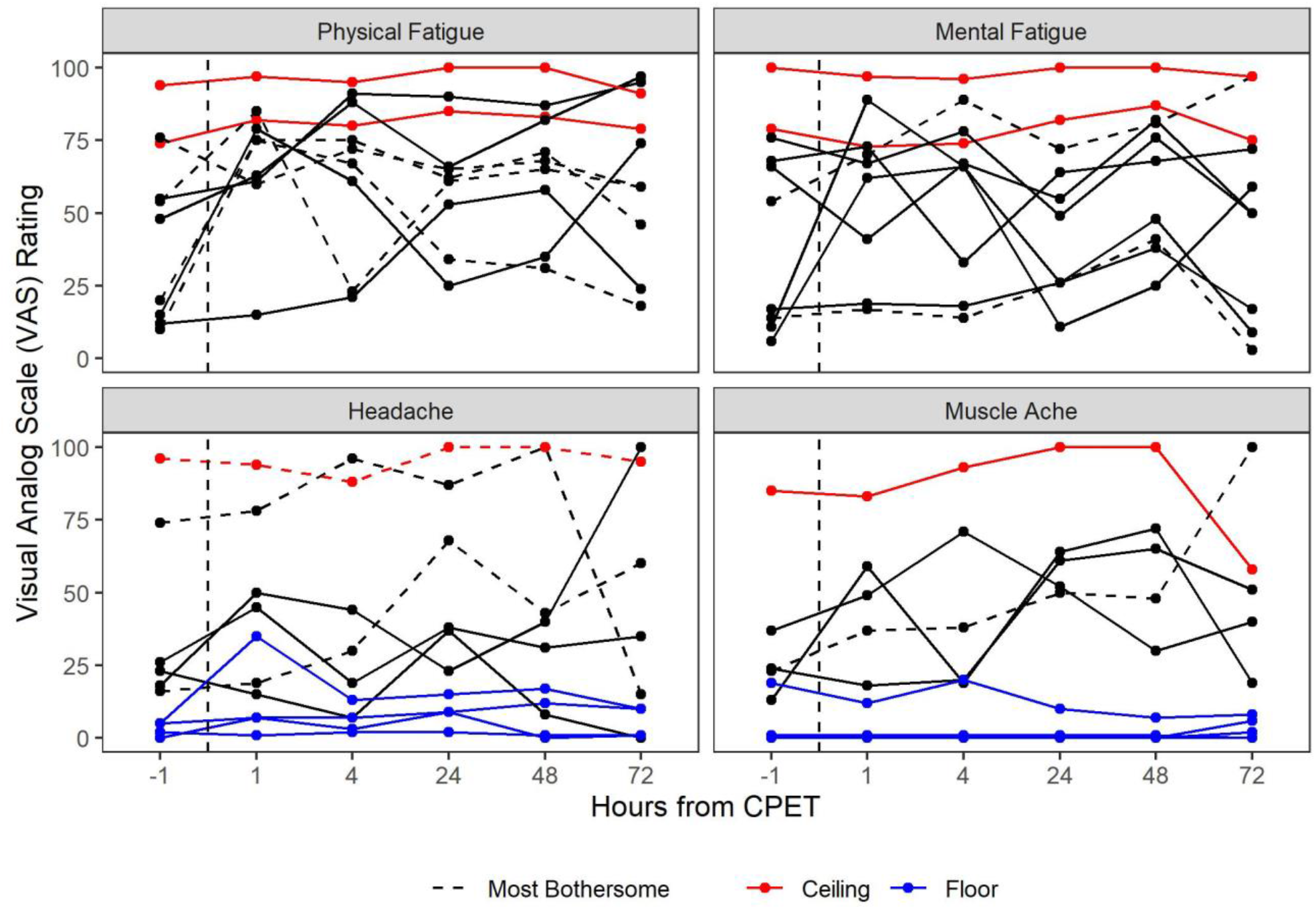
Visual Analog Scale Data for ME/CFS patients (n=10).

## DISCUSSION

ME/CFS has been described as a devastating disabling illness (4, 38) and CPET is an important tool for measuring PEM, yet the lack of a gold standard method for the assessment of PEM following CPET hinders research in the field. This is the first to utilize a mixed methods approach to assess the experience of PEM in ME/CFS and healthy volunteers via QIs and compare it to the traditional VAS questionnaire method. The VAS effectively captured the absence of PEM in HVs but did not capture the nuanced experienced of PEM in 30% of ME/CFS volunteers. The addition of QIs enabled determination of a PEM peak in all of the ME/CFS volunteers.

A wide variation was seen across individuals. Several symptom scores remained high or low throughout the six timepoints, demonstrating the potential for VAS to produce ceiling and floor effects. The QI approach allowed for the determination of PEM peaks, captured the most bothersome symptom experienced, and provided a more granular measurement even when VAS scores were flat. While floor and ceiling effects in VAS are not unique to ME/CFS (39, 40), the high burden of premorbid symptoms makes the VAS uniquely susceptible to these effects when used serially. Further, the correlations between QIs and VAS scales were relatively strong at baseline but became serially weaker at time points after exercise, again suggesting that VAS scales are less sensitive to change than QIs. It appears performance of VAS scales can be improved by use of QI measurements.

While fatigue is the most common symptom associated with PEM in the literature (20, 31, 4), it was the most bothersome symptom for only 40% of the ME/CFS volunteers in this study.

Perhaps this explains why physical fatigue severity VAS did not capture PEM for half of ME/CFS volunteers. Performance was improved when most bothersome symptoms were considered, with strong correlations between QIs and VAS seen in seven of ten ME/CFS volunteers. However, for the other three volunteers, the correspondence was low and the most bothersome VAS still would not be able to capture the occurrence of PEM. This suggests that any singular quantitative VAS measure cannot effectively capture the multidimensional experience of PEM.

Numerous ME/CFS studies have documented that the experience 10+ PEM symptoms (4, 20, 31) both following CPET and in day-to-day experience is common. Due to the wide breadth of symptoms, high levels of pain, and other symptoms experienced during PEM (37, 20, 10), a more nuanced measurement system with an ability to consider multiple symptoms as a single holistic experience may be necessary to accurately access PEM. While all ME/CFS volunteers experienced fatigue, the QI data remind that each person’s manifestation of PEM is unique. One volunteer was observed curled up in the fetal position with extreme sensory discomfort, while another twitched uncontrollably with heightened anxiety. Perhaps the focus of measuring PEM should be on the totality of disability rather than any individual symptom.

Future research is required to determine the utility of the addition of a most bothersome symptom assessment to traditional VAS measures. It is not clear if the most bothersome symptom can be adequately collected retrospectively. It was noted that for a quarter of ME/CFS volunteers, the most bothersome symptom could change across the different time points when queried. In this work, a single most bothersome symptom was determined by team consensus after an analysis of the patient’s textual descriptions of symptoms. Other approaches that allow for most bothersome symptoms to change from timepoint to timepoint may work better than what was presented in the current study. These results also suggest that categorical scales (e.g. “a little better,” “a lot better,” “a little worse,” “a lot worse”) may also be considered in lieu of VAS scales for measuring PEM. These alternative approaches need exploration and testing to improve upon the ability to measure individual PEM experiences.

## STUDY LIMIATIONS

As with all studies, this study has limitations. First, the current study has a small sample size. However, the sample size was adequate to demonstrate the added value of using QIs in conjunction with VAS for assessment of PEM and demonstrate the deficiencies of standard methodologies. A larger cohort would allow for a more accurate estimate of the added value and deficiency. Second, neither QIs nor the VAS have been validated to assess PEM following CPET. Nevertheless, the current study used rigorous qualitative methods and analyzed each patient’s unique experience of PEM in a way not possible with VAS scales used in isolation.

Future studies could assess this methodology using respondent validation. The use of QIs to assess PEM requires more time and resources than questionnaires alone, which impacts the implementation of this methodology.

## CONCLUSION

Determining best practices for the measurement of PEM following CPET will enable a better understanding of this disabling condition. Comparing severity data collected from QIs and VAS in the current study revealed potential methods to address some of the known limitations of the VAS in capturing PEM following CPET. QIs allowed for synthesizing the entire experience of PEM through probing and clarifying that is not possible with quantitative or open-ended questionnaires and outperformed VAS scales in determining the onset of PEM. Future research with a larger sample size should use QIs to validate the approach and further explore how best to combine QIs and VAS measures for optimal assessments of PEM in ME/CFS and Long COVID.

## Data Availability

All data produced in the present study are available upon reasonable request, pending review for confidentiality.

## ACKNOWLEDGEMENTS

We would like to thank Drs. Mark Hallett, Leorey Saligan and Termeh Feinberg for helpful comments on a previous draft. We would also like to thank the entire NIH ME/CFS Study Group.

## AUTHOR CONTRIBUTIONS

BS designed the study, collected and analyzed the data, wrote and edited the manuscript. BC AG and SC collected and analyzed the data and edited the manuscript. GN analyzed the data, participated in writing and editing the paper and approved of the final version. AN contributed to supervision of the work, participated in writing and editing the paper, and approved the final version. BW contributed to supervision of the work, data collection, data analysis, writing and editing the paper, and approved the final version.

## ABBREVIATIONS

PEM: post-exertional malaise
QI: qualitative Interview
ME/CFS: Myalgic Encephalomyelitis/Chronic Fatigue Syndrome
VAS: visual analog scale
CPET: cardiopulmonary exercise test
NRS: numeric rating scale

## Notes

### Competing Interest Statement

The authors have declared no competing interest.

### Clinical Trial

NCT02669212

### Funding Statement

This research was supported (in part) by the Intramural Research Program of the NIH, National Center for Complementary and Integrative Health, and the National Institute of Neurological Diseases and Stroke.

### Author Declarations

This study was carried out in accordance with the recommendations of the NIH Institutional Review Board (IRB), NIH with written informed consent from all subjects. The protocol was approved by the NIH IRB.

